# Structural and metabolic brain abnormalities in COVID-19 patients with sudden loss of smell

**DOI:** 10.1101/2020.10.18.20214221

**Authors:** Maxime Niesen, Nicola Trotta, Antoine Noel, Tim Coolen, Georges Fayad, Gil Leurkin-Sterk, Isabelle Delpierre, Sophie Henrard, Niloufar Sadeghi, Jean-Christophe Goffard, Serge Goldman, Xavier De Tiège

**Author notes:** Corresponding Author: Maxime Niesen, Laboratoire de Cartographie fonctionnelle du Cerveau, UNI - ULB Neurosciences Institute, Université libre de Bruxelles (ULB), 1070 Brussels, Belgium. Conflict of interest disclosure: None reported. Role of the Funder/Sponsor: The funding sources had no role in the design and conduct of the study (collection, management, analysis, and interpretation of data); in the preparation, review and approval of the manuscript; and decision to submit the manuscript for publication. Data availability: De-identified brain MRI and FDG-PET data can be shared upon reasonable request for scientific purpose and after approval of the CUB Hôpital Erasme Ethics Committee and authorities. Standard protocol approvals, registrations, and patient consents: Studies were carried out at the CUB Hôpital Erasme (Brussels, Belgium). Participants contributed to the corresponding studies approved by the institutional Ethics Committee after written informed consent (References: P2020/204, CCB: B4062020000031; P2017/541, CCB: B406201734197).

## Abstract

**Objectives:** Sudden loss of smell is a very common symptom of coronavirus disease 19 (COVID-19). This study characterizes the structural and metabolic cerebral correlates of dysosmia in patients with COVID-19.

**Methods:** Structural brain magnetic resonance imaging (MRI) and positron emission tomography with [18F]-fluorodeoxyglucose (FDG-PET) were prospectively acquired simultaneously on a hybrid PET-MR in twelve patients (2 males, 10 females, mean age: 42.6 years, age range: 23-60 years) with sudden dysosmia and positive detection of severe acute respiratory syndrome coronavirus 2 (SARS-CoV-2) on nasopharyngeal swab specimens. FDG-PET data were analysed using a voxel-based approach and compared with that of a group of healthy subjects.

**Results:** Bilateral blocking of the olfactory cleft was observed in six patients, while subtle olfactory bulb asymmetry was found in three patients. No MRI signal abnormality downstream of the olfactory tract was observed. Heterogeneous (decrease or increase) glucose metabolism abnormalities were observed in core olfactory and high-order neocortical areas. A modulation of regional cerebral glucose metabolism by the severity and the duration of COVID-19-related dysosmia was disclosed using correlation analyses.

**Conclusions:** This PET-MR study shows that sudden loss of smell in COVID-19 is not related to central involvement due to SARS-CoV-2 neuroinvasiveness. Loss of smell is associated with heterogeneous cerebral metabolic changes in core olfactory and high-order cortical areas likely related to combined processes of deafferentation and active functional reorganisation secondary to the lack of olfactory stimulation.

## Introduction

The severe acute respiratory syndrome coronavirus 2 (SARS-CoV-2) is responsible, since December 2019, of an ongoing worldwide outbreak of atypical and severe pneumonia, called coronavirus disease 2019 (COVID-19). Sudden loss of smell (i.e., anosmia) and taste (i.e., ageusia) are very common symptoms in SARS-CoV-2 infection, being reported in up to 80% of cases (for a review, see [1]).

SARS-CoV-2 infects human cells by binding to the angiotensin-converting enzyme 2 (ACE2) with contribution of the transmembrane protease serine 2 (TMPRSS2).[2] ACE2 and TMPRSS2 appear to be widely expressed by key support (sustentacular cells, Bowman’s gland, microvillar cells) and stem cells of the olfactory epithelium (OE), but not by olfactory sensory neurons (OSNs).[1] How SARS-CoV-2 infection of OE cells leads to sudden smell loss remains hypothetical. The causal link might involve (i) blockage of the olfactory cleft due to focal OE inflammation, (ii) alteration of OSNs function, or (iii) widespread damage to OE and subsequently OSNs with variable time to recovery depending on the extent of stem cells damage.[1] Changes in olfactory bulbs (OB) and variable cortical (e.g., orbitofrontal cortex) or subcortical (e.g., thalamus, hypothalamus, brainstem) areas have also been disclosed by magnetic resonance imaging (MRI)[3–7] or positron emission tomography (PET)[8,9] in some COVID-19 patients with or without anosmia. Whether these central nervous system (CNS) abnormalities are indicative of potential SARS-CoV-2 neuroinvasion triggering anosmia is still a matter of debate.[1]

To get novel insights into the possible CNS involvement in COVID-19 patients with sudden anosmia, we used a hybrid PET and MRI system (PET-MR) to investigate the brain structure and regional glucose metabolism of SARS-CoV-2-infected patients with sudden loss of smell. We specifically searched for MRI signal and metabolic abnormalities along the olfactory system and brain areas previously shown to be involved in the pathophysiology of non-COVID-19-related anosmia. We also searched for any sign of acute brain abnormalities.

## Methods

### Study design and participants

From April 2020 to the end of May 2020, dysosmic patients in the context of a SARS-Cov-2 infection were prospectively recruited at the CUB Hôpital Erasme based on the following criteria: (i) sudden loss of smell; (ii) positive SARS-CoV-2 detection (direct antigen detection or reverse transcriptase polymerase chain reaction) on nasopharyngeal swab specimen; (iii) no prior history of neurological, psychiatric, nasal, sinus or olfactory disorders; (iv) MRI compatibility; (v) formal acceptance to participate to the study.

Objective evaluation of olfactory dysfunction was performed using the “*identification test*” included in the Sniffin Sticks Battery (Burghart Messtechnik, Wedel, Germany) at the time of PET-MR data acquisition. The evolution of dysosmia was assessed at the time of manuscript submission using qualitative anamnesis and visual analog scale (0, no smell at all; 10, normal olfactory function). Clinical data were retrieved from patients’ anamnesis and medical records.

For PET data analyses, a PET-MR data set acquired in the context of other neuroimaging studies and composed of twenty-six healthy subjects (5 females, 21 males, mean age: 35 years, age range: 22-52 years) without any history of COVID-19, smell or taste disorders was used as control group.

### Standard protocol approvals, registrations, and patient consents

Studies were carried out at the CUB Hôpital Erasme (Brussels, Belgium). Participants contributed to the corresponding studies approved by the institutional Ethics Committee after written informed consent (References: P2020/204, CCB: B4062020000031; P2017/541, CCB: B406201734197).

### PET-MR data acquisition

Cerebral PET and MRI data were obtained using a 3T hybrid PET-MR scanner (SIGNA™, GE Healthcare, Chicago, IL) in accordance with COVID-19 institutional hygienic and safety rules.

MRI sequences have been described in details elsewhere.[5] They consisted in whole-brain axial 3D T1-weighted imaging (WI), axial T2WI, sagittal 3D T2WI fluid-attenuated inversion recovery (FLAIR), axial 3D susceptibility-WI (SWI), axial diffusion-WI (DWI), and a coronal T2WI centered on olfactory bulbs. For PET data acquisitions, participants fasted for at least 4 hours, were awake in an eye-closed rest and received an intravenous bolus injection of 3-5 mCi (111-185 MBq) of [18F]-fluorodeoxyglucose (FDG) before PET-MR data acquisition. Forty minutes post-injection, a 20-min data acquisition was performed. PET data were reconstructed using the fully 3D iterative reconstruction algorithm VUE Point FX-S, which takes into account the time-of-flight (TOF) information and the correction for the point spread function (PSF) of the system. The algorithm was configured with 10 iterations, 28 subsets and a standard Z-axis filter cut-off at 4 mm. The photons’ attenuation was corrected with a MRI-based map (MRAC) acquired simultaneously. PET images were displayed in a 256 × 256 x 89 matrix format, with a slice thickness of 2.78 mm. The reconstructed files were downloaded in their original format (DICOM, ECAT, Interfile) for meta-information and converted in NIfTI format for subsequent analysis.

### MRI data analysis

MRI data were reviewed by two experienced neuroradiologists (T.C. and I.D.) following a systematic and comprehensive visual assessment procedure previously described.[5] Brain MRI findings were divided into recent (i.e., potentially related to SARS-CoV-2) and long-standing (i.e., unlikely related to SARS-CoV-2) changes.[5] The Lund-Mackay score for chronic rhinosinusitis[10] was used for the visual assessment of incidental paranasal sinus inflammation and adapted to evaluate olfactory cleft obliteration (0: normal; 1: partial obliteration; 2: total obliteration) at the level of the OB. Final reports were then discussed with neurologists (S.G., X.D.T.), neuroradiologist (N.S.), otorhinolaryngologists (M.N., A.N., G.F.), and internists (S.H., J-C.G.)

### PET data analysis

FDG-PET data were analyzed using the voxel-based Statistical Parametric Mapping software (SPM8, http://www.fil.ion.ucl.ac.uk/spm/, Wellcome Trust Centre for Neuroimaging, London, UK) based on conventional subtractive and correlation analyses previously used by our group and described in details elsewhere.[11–15]

Briefly, PET images were spatially normalized into the Montreal Neurologic Institute template (MNI, Montreal Neurologic Institute, Quebec, Canada) and then smoothed using a 12-mm full-width at half-maximum Gaussian isotropic kernel. Global activity normalization was performed by proportional scaling.

For individual-and group-level subtractive analyses, we constructed general linear models (GLMs) of the preprocessed FDG-PET data of dysosmic patients —taken individually or as a group— and healthy subjects taken as separate groups. Separate *t*-contrasts identified brain areas where glucose metabolism was significantly lower or higher in dysosmic patient(s) compared to healthy subjects.

For correlation analyses, we constructed GLMs of the preprocessed FDG-PET data of dysosmic patients taken as one group. Scores of the *identification test* (Sniffin Sticks Battery) and duration (days) of smell loss were introduced as covariates of interest centered around condition means in separate GLMs. Separate *t*-contrasts identified brain regions showing significant positive or negative correlations between the covariates of interest and regional cerebral glucose metabolism. For significant voxels, regression plots and associated slope coefficients/p-values (Pearson’s correlation) between glucose metabolism and covariates of interest were obtained in Matlab R2017a (MathWorks Inc.).

Participant’s age were introduced as covariate of no-interest in all GLMs to avoid potential ageing confounds. Gender was not used as covariate of no-interest considering the unbalanced gender profile between dysosmic patients and healthy subjects.

Results were considered significant at *p* < .05 corrected for multiple comparisons (Family Wise Error, FWE). To minimize type II errors, results were also deemed significant at *p* < .001 uncorrected (cluster size *k* ≥ 50 voxels) if voxels involved brain areas previously reported as (i) involved in the processing of odors in healthy subjects[16–19], or as (ii) metabolically abnormal in COVID-19-related or non-COVID-19-related anosmia.[8,9,20–22]

## Results

### Patients’ clinical characteristics

Table 1 describes the patients’ clinical characteristics.

**Table 1:** Clinical characteristics of the patients

| Clinical characteristic | % of Patients ($n_{tot} = 12$) |
| --- | --- |
| **Mean age (range), years** | 42.6 (23 – 60) |
| **Mean Body Mass Index (range), kg/m<sup>2</sup>** | 24.8 (22 – 29.8) |
| **Female gender** | 83.3 (10) |
| **Active smoker** | 8.3 (1) |
| **Hospitalisation** | 8.3 (1) |
| **SARS-CoV-2 detection** |  |
| Direct antigen detection | 25 (3) |
| Reverse transcriptase PCR | 75 (9) |
| **Comorbidities** |  |
| Respiratory allergy | 50 (6) |
| Asthma | 8.3 (1) |
| Hypertension | 16.7 (2) |
| Hypothyroidism | 8.3 (1) |
| Diabetes | 0 |
| **Symptoms** |  |
| Dysosmia | 100 (12) |
| Dysgueusia | 91.7 (11) |
| Myalgia, Arthralgia | 75 (10) |
| Cough | 66.7 (8) |
| Rhinorrhea | 66.7 (8) |
| Fever | 50 (6) |
| Headache | 50 (6) |
| Dyspnea | 41.7 (5) |
| Asthenia | 41.7 (5) |
| Diarrhea | 25 (3) |
| Nausea, Vomiting | 16.7 (2) |
| Rash | 8.3 (1) |
| Throat pain | 8.3 (1) |
| **Objective olfactory assessment** |  |
| Anosmia | 58.3 (7) |
| Hyposmia | 41.7 (5) |

Fourteen patients (11 females, 3 males, mean age: 43 years, age range: 23–60 years) fulfilled the inclusion criteria. Two patients were excluded from further analyses due to magnetic artifacts covering the nasal cavity and orbitofrontal cortex (1 patient), and to the presence of a large right sylvian arachnoid cyst that would interfere with the normalization procedure (1 patient). Therefore, the final sample considered for the study was composed of 12 patients.

In those patients, subjective loss of smell occurred on average 15 days (+/- 9 days) before PET-MR data acquisition. It was the predominant symptom related to SARS-CoV-2 infection in seven patients. Other symptoms were dyspnea (five patients), asthenia (five patients) and gastro-intestinal symptoms (three patients). Six patients also presented with headaches. One patient was hospitalized for the management of COVID-19 respiratory symptoms, while the others were outpatients.

Olfactory disorder was confirmed in each of the twelve SARS-CoV-2-positive patients by objective olfactory testing. The mean *identification test* score was 8/16 (+/- 1.4), and documented hyposmia in 5 patients (scores between 9 and 11) and anosmia in 7 patients (scores ≤ 8).

Positive evolution of dysosmia was observed in all patients. Five patients fully or almost fully recovered (4 to 10 weeks after onset; visual analog scale: 9 or 10). Seven patients still complained of relative hyposmia (improvement observed from 1 to 16 weeks after onset; visual analog scale: 6 to 8)

### Structural MRI abnormalities

Table 2 and Figure 1 describe the MRI findings observed in dysosmic patients.

**Table 2:**
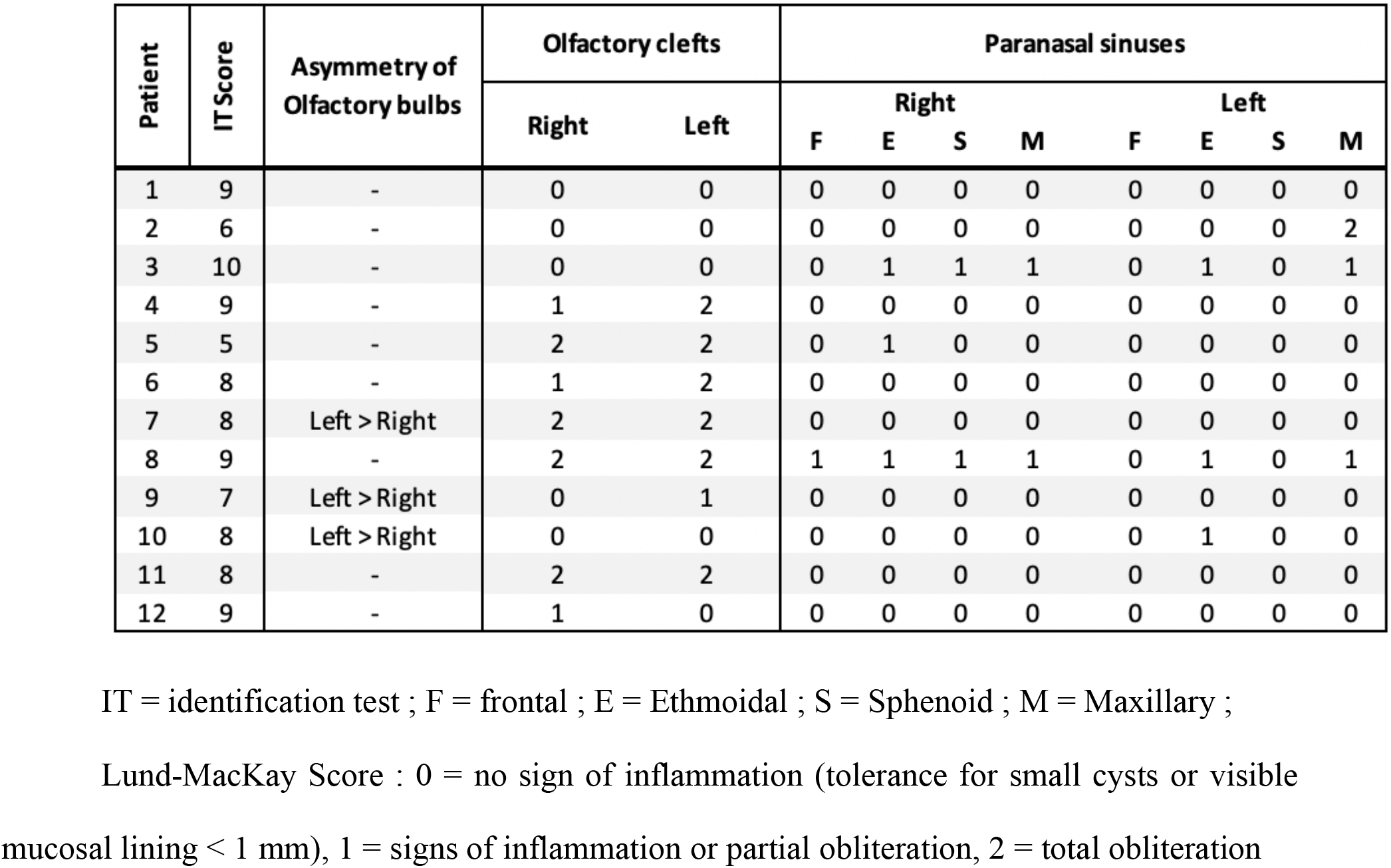
*Structural* MRI Findings

| Patient | IT Score | Asymmetry of Olfactory bulbs | Olfactory clefts |  | Paranasal sinuses |  |  |  |  |  |  |  |
| --- | --- | --- | --- | --- | --- | --- | --- | --- | --- | --- | --- | --- |
|  |  |  | Right | Left | Right |  |  |  | Left |  |  |  |
|  |  |  |  |  | F | E | S | M | F | E | S | M |
| 1 | 9 | - | 0 | 0 | 0 | 0 | 0 | 0 | 0 | 0 | 0 | 0 |
| 2 | 6 | - | 0 | 0 | 0 | 0 | 0 | 0 | 0 | 0 | 0 | 2 |
| 3 | 10 | - | 0 | 0 | 0 | 1 | 1 | 1 | 0 | 1 | 0 | 1 |
| 4 | 9 | - | 1 | 2 | 0 | 0 | 0 | 0 | 0 | 0 | 0 | 0 |
| 5 | 5 | - | 2 | 2 | 0 | 1 | 0 | 0 | 0 | 0 | 0 | 0 |
| 6 | 8 | - | 1 | 2 | 0 | 0 | 0 | 0 | 0 | 0 | 0 | 0 |
| 7 | 8 | Left > Right | 2 | 2 | 0 | 0 | 0 | 0 | 0 | 0 | 0 | 0 |
| 8 | 9 | - | 2 | 2 | 1 | 1 | 1 | 1 | 0 | 1 | 0 | 1 |
| 9 | 7 | Left > Right | 0 | 1 | 0 | 0 | 0 | 0 | 0 | 0 | 0 | 0 |
| 10 | 8 | Left > Right | 0 | 0 | 0 | 0 | 0 | 0 | 0 | 1 | 0 | 0 |
| 11 | 8 | - | 2 | 2 | 0 | 0 | 0 | 0 | 0 | 0 | 0 | 0 |
| 12 | 9 | - | 1 | 0 | 0 | 0 | 0 | 0 | 0 | 0 | 0 | 0 |

IT = identification test ; F = frontal ; E = Ethmoidal ; S = Sphenoid ; M = Maxillary ;
Lund-MacKay Score : 0 = no sign of inflammation (tolerance for small cysts or visible mucosal lining < 1 mm), 1 = signs of inflammation or partial obliteration, 2 = total obliteration

**Figure 1:**
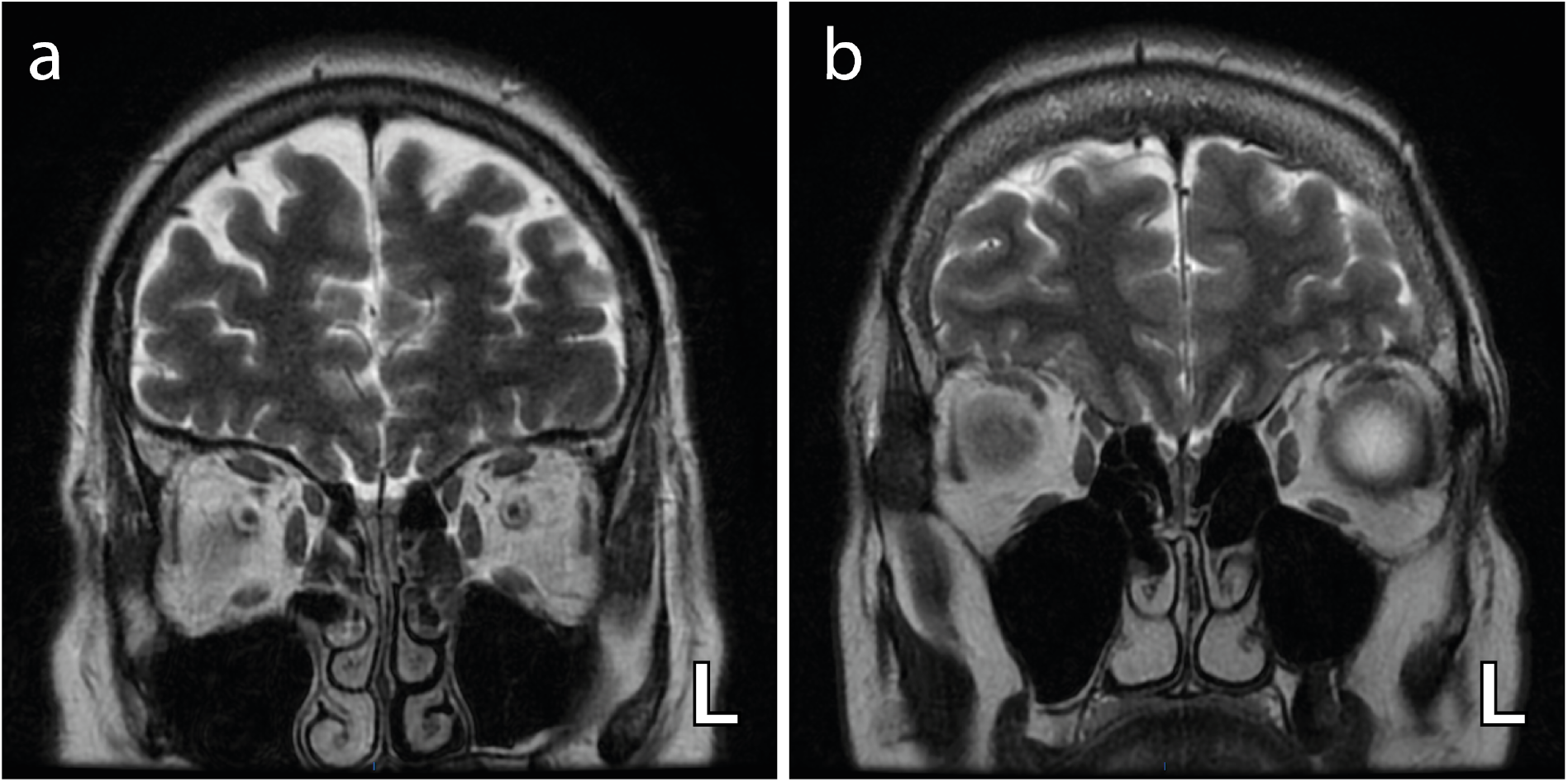
Axial T2-weighted coronal images demonstrating bilateral and complete obliteration of the olfactory clefts (**a**) with no associated olfactory bulb asymmetry and (**b**) with asymmetry of the olfactory bulbs (left (L) bulb relatively enlarged).

Subtle OB asymmetry was found in three patients with no associated bulbar signal abnormality. Four patients had no olfactory cleft obliteration, two had partial obliteration of one olfactory cleft, and six patients had bilateral olfactory cleft obliteration (bilateral and complete in four). Discrete signs of sinus inflammation were found in six patients (Lund-Mackay score less than 6). No sign of recent brain abnormality was found in any of the patients. Non-systematic long-standing brain MRI findings (e.g., periventricular halo, small arterial aneurysm, developmental venous anomaly, focal microbleed) were found in all but one patient. Of note, we did not find any significant correlation between the *identification test* score and the Lund-Mackay score of the olfactory cleft (Pearson correlation, *r* = –0.178, *p* = 0.58) or with the Lund-Mackay score of the paranasal sinuses (Pearson correlation, *r* = 0.276, *p* = 0.39).

### Regional cerebral glucose abnormalities

Tables 3-4 and Figures 2-4 describe the regional cerebral glucose abnormalities observed at the individual (Figure 2) and group (Figure 3) levels in dysosmic patients, as well as the results of correlation analyses (Figure 4).

**Table 3:**
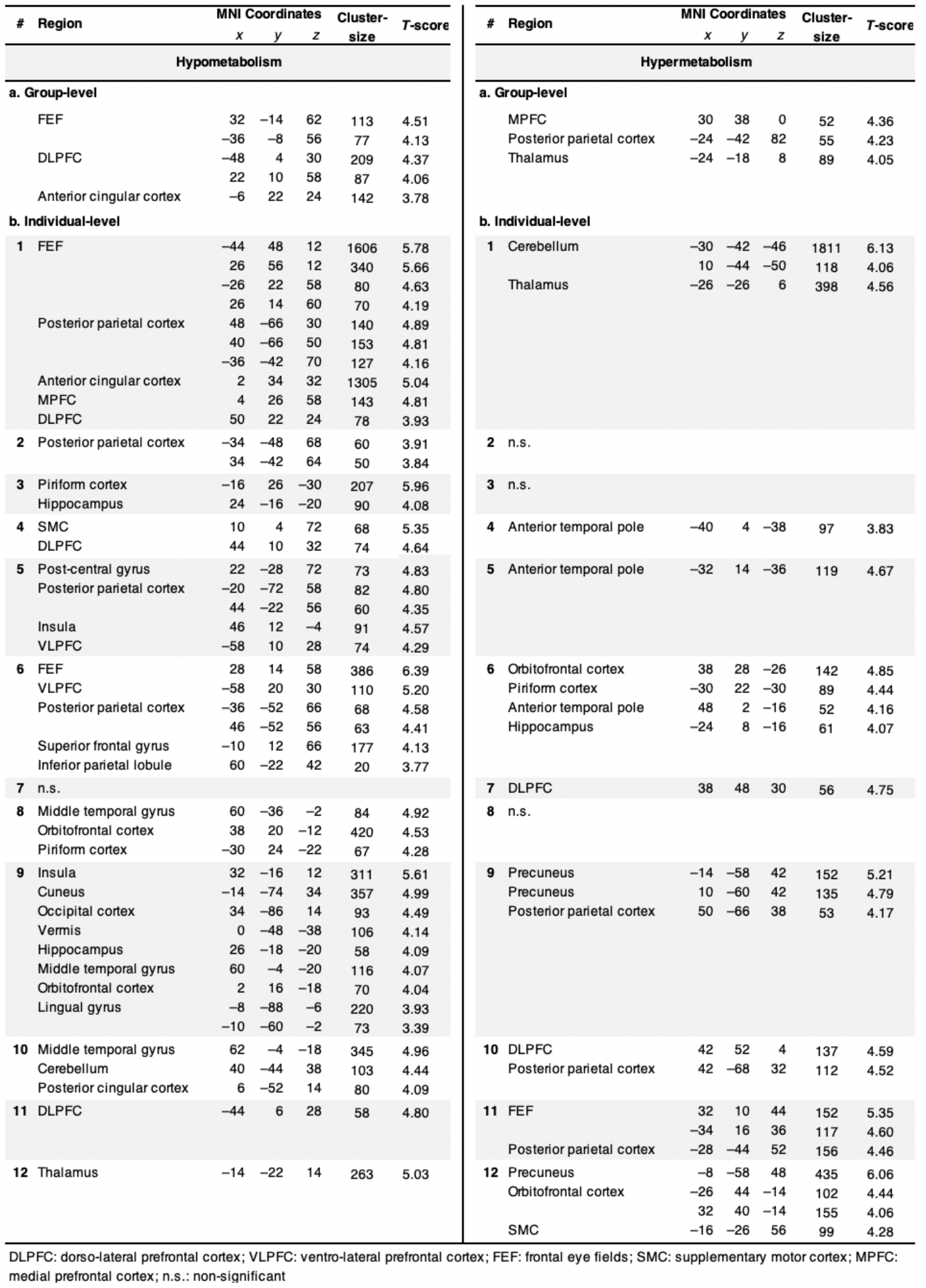
MNI coordinates, cluster size (k) and t-value (T) of cortical areas with significant hypo- and hypermetabolism (**a**) at the group-level, and (**b**) at the individual-level.

**Table 4:** MNI coordinates, cluster size (k), Pearson’s correlation coefficient (*r*) and T-score of cortical areas with significant correlation between the olfactory function (assessed by the *Identification test* score) and cerebral glucose metabolism; and between the duration of smell loss and brain metabolism.

| Covariate of interest | Region | MNI Coordinates |  |  | Cluster-size | Pearson's *r* | *T*-score |
| --- | --- | --- | --- | --- | --- | --- | --- |
|  |  | *x* | *y* | *z* |  |  |  |
| Olfactory function | FEF | -30 | 6 | 48 | 87 | -0.97 | 12.60 |
|  | Posterior parietal cortex | 22 | -54 | 46 | 60 | -0.94 | 8.20 |
|  | Orbitofrontal cortex | 12 | 68 | 0 | 88 | -0.92 | 7.07 |
|  |  | 38 | 54 | -10 | 61 | -0.92 | 6.68 |
| Duration of anosmia | MPFC | 0 | 48 | 46 | 113 | 0.96 | 10.25 |
|  |  | -6 | 32 | 62 | 68 | 0.93 | 8.08 |
|  | DLPFC | -38 | 32 | 34 | 143 | 0.94 | 8.73 |
|  |  | -48 | 24 | 18 | 87 | 0.90 | 6.32 |
|  | Orbitofrontal cortex | 28 | 66 | 0 | 50 | 0.91 | 6.57 |
|  |  | -34 | 32 | -10 | 66 | 0.90 | 6.44 |
|  |  | 36 | 34 | -14 | 198 | 0.89 | 5.97 |
|  | Posterior parietal cortex | -24 | -66 | 54 | 73 | 0.90 | 6.25 |

DLPFC: dorso-lateral prefrontal cortex; FEF: frontal eye fields; MPFC: medial prefrontal cortex

**Figure 2:**
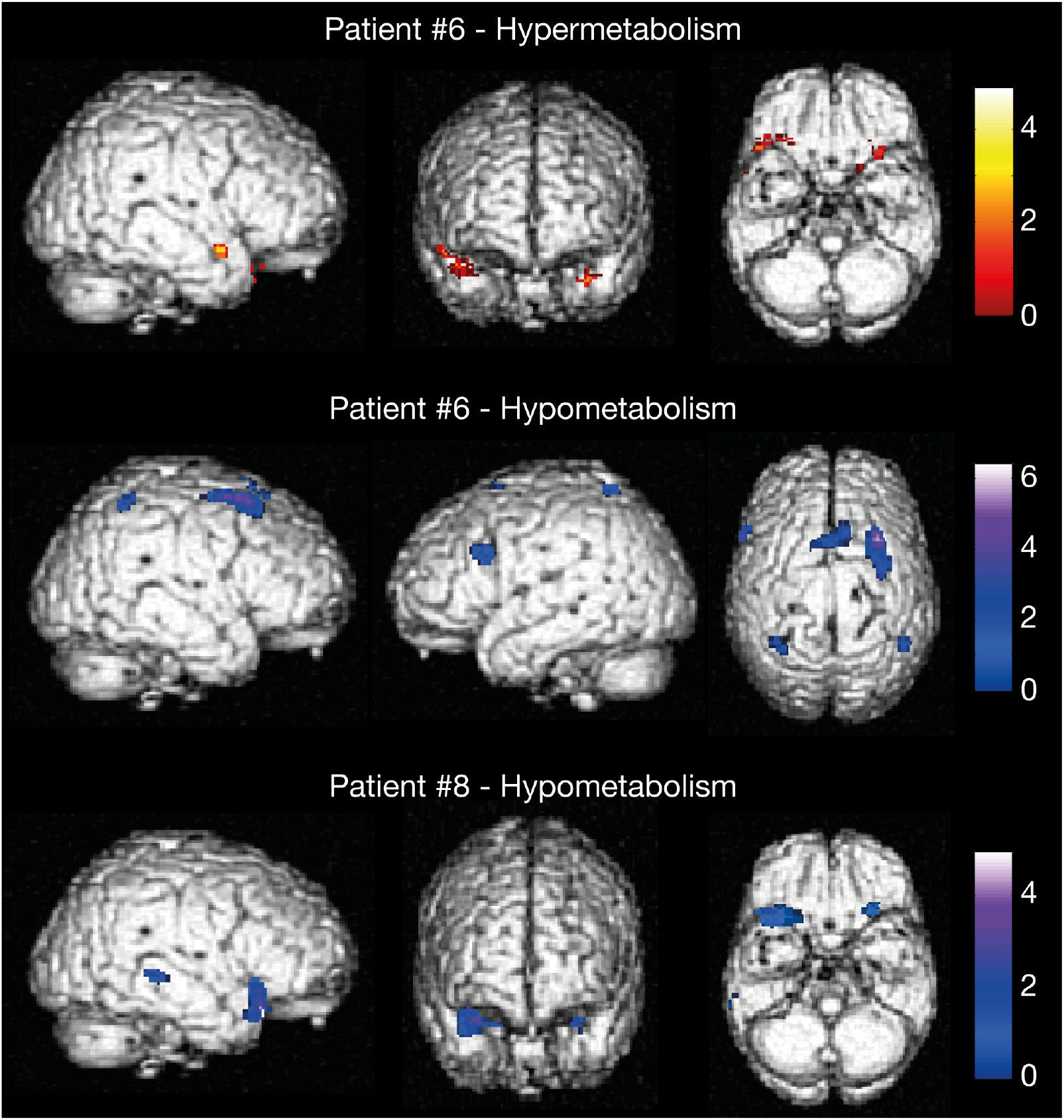
Regional hyper- and hypometabolism in two typical patients. Legend: Images are thresholded at *p* < .001 uncorrected (k ≥ 50 voxels).

**Figure 3:**
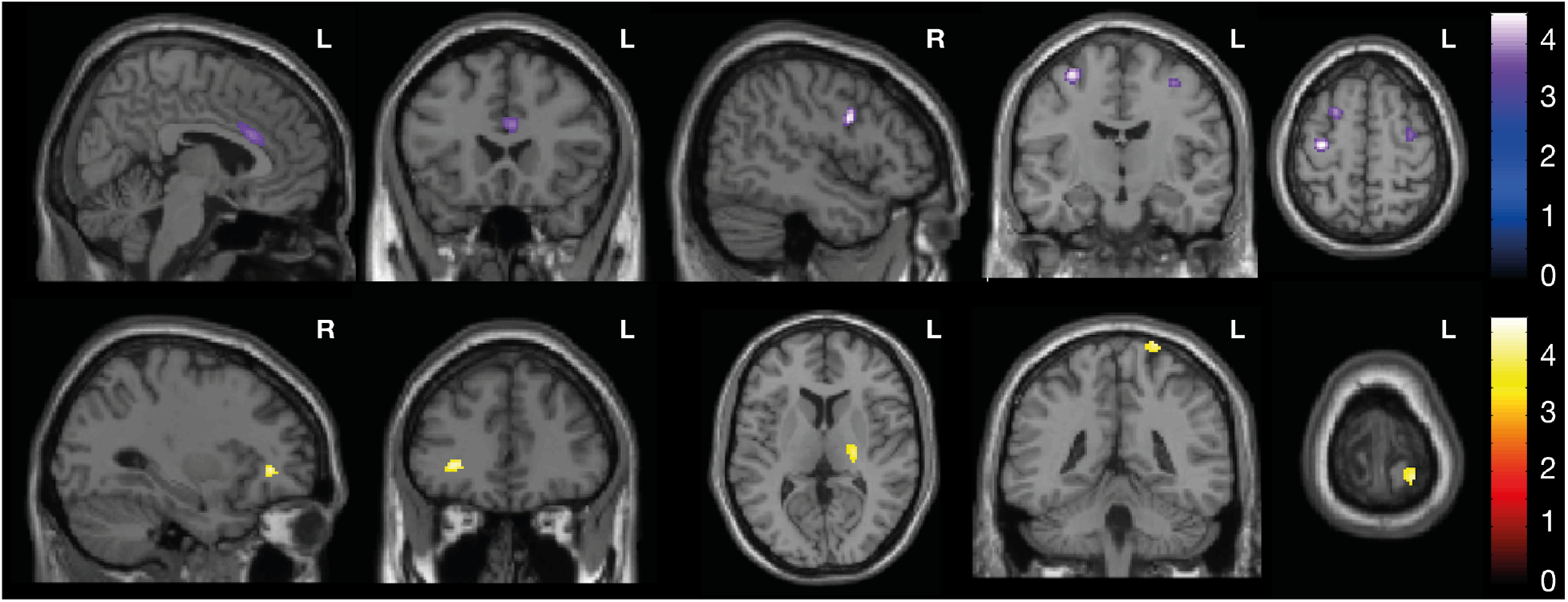
Regional cerebral glucose metabolic abnormalities observed at the group-level. Legend: Regional hypometabolism (Top) and hypermetabolism (Bottom) observed at the group level. Images are thresholded at *p* `< .001 uncorrected (k ≥` 50 voxels).

**Figure 4:**
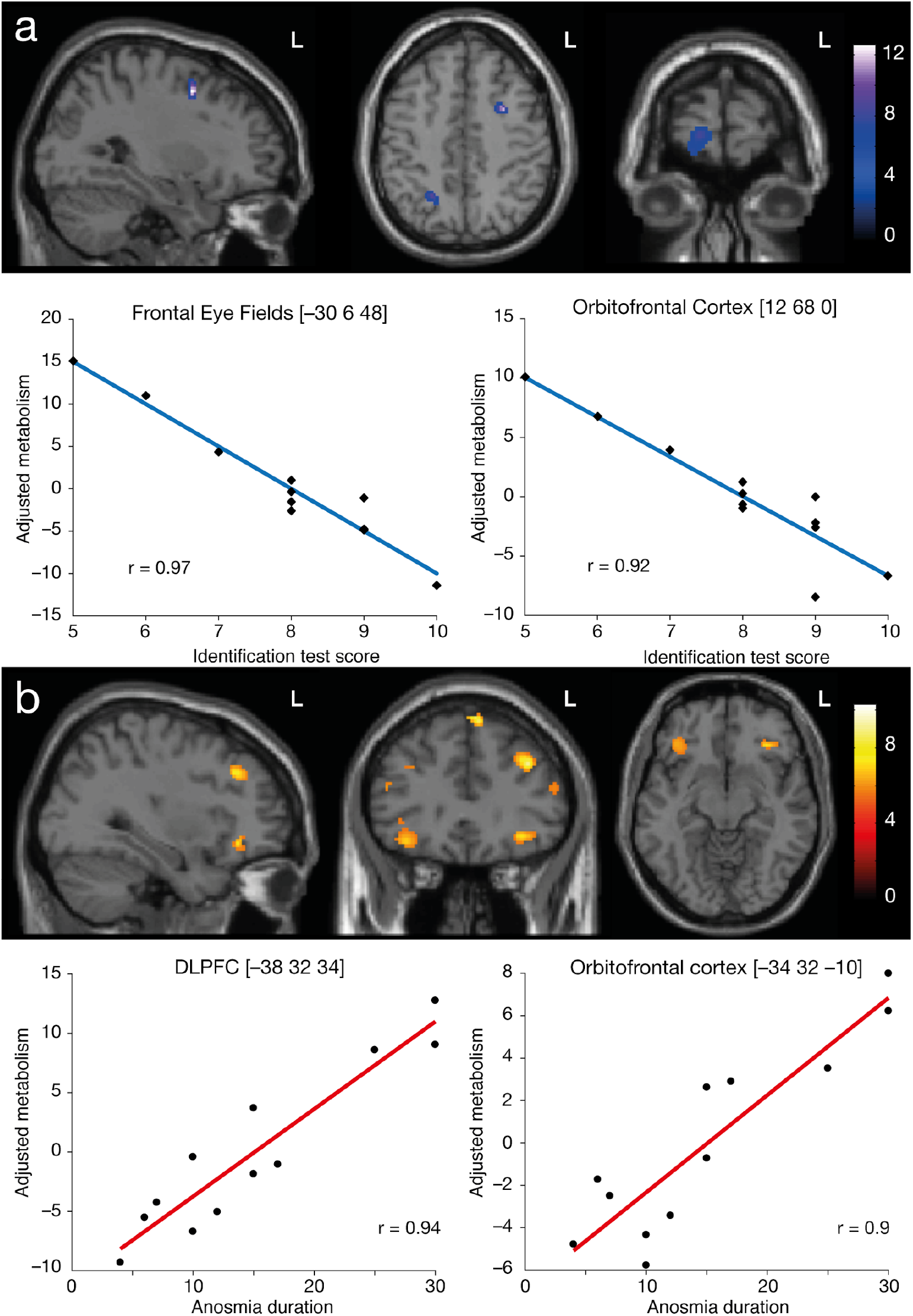
Results of the correlation analyses performed between the cerebral glucose metabolism and the severity of olfactory dysfunction, and between the cerebral glucose metabolism and the duration of anosmia. (**a**) Regression plots of the severity of olfactory dysfunction (assessed by *the identification test score*) and adjusted metabolic responses obtained by considering the peak voxel in the left FEF ([–30 6 48], Pearson’s correlation: *r* = 0.97, *p* < .001) and in the right orbitofrontal cortex ([12 68 0], Pearson’s correlation: *r* = 0.94, *p* < .001). (**b**) Regression plots of the duration of anosmia and adjusted metabolic responses obtained by considering the peak voxel in the left DLPFC ([–38 32 34], Pearson’s correlation: *r* = 0.94, *p* < .001) and in the left orbitofrontal cortex ([–34 32 –10], Pearson’s correlation: *r* = 0.9, *p* < .001).

At the individual-level, all patients showed significant (*p* < .001 uncorrected) decrease (3 patients) or increase (1 patient) in regional cerebral glucose metabolism, or a combination of both (8 patients). Hypometabolic areas involved nodes of the core olfactory network (4 patients), somatosensory (1 patient) or visual (1 patient) cortical areas, the cerebellum (2 patients), or higher-order neocortical areas (9 patients) mainly encompassing the attentional, emotional or default mode (DMN) networks. Hypermetabolic areas also involved nodes of the core olfactory network (6 patients), the cerebellum (1 patient), or high-order neocortical areas (5 patients) belonging to the attentional network or the DMN.

At the group-level, compared with healthy controls, dysosmic patients presented significant (*p* < .001 uncorrected) decrease in regional glucose consumption in bilateral dorso-lateral prefrontal cortex (DLPFC), bilateral frontal eye fields (FEF), and in the left anterior cingular cortex (ACC). Significant (*p* < .001 uncorrected) hypermetabolic areas were also found in the right medial prefrontal cortex (MPFC), the left posterior parietal cortex (PCC) and in the left thalamus.

Significant (*p* < .001 uncorrected) negative correlation was found between the *Identification test* score and cerebral glucose metabolism in the left FEF, the right orbitofrontal cortex and in the right PCC, indicating that a more severe deficit was related to a higher glucose metabolism in these brain regions.

Significant (*p* < .001 uncorrected) positive correlation was found between the duration of dysosmia and brain metabolism in bilateral orbitofrontal cortex, and in the left DLPFC, MPFC and PPC, indicating that a longer duration of loss of smell was related to a higher glucose metabolism in these brain regions.

Of note, no significant correlation was found between the *Identification test* score and the duration of dysosmia (Pearson correlation, *r* = 0.07, *p* = 0.82).

## Discussion

This PET-MR study performed in twelve patients with sudden loss of smell due to SARS-CoV-2 infection demonstrates (i) bilateral obliteration of the olfactory cleft in 50% of the patients, (ii) subtle OB asymmetry in 25% of the patients, (iii) the absence of MRI signal abnormality downstream of the olfactory tract in all patients, (iv) heterogeneous glucose metabolism abnormalities mainly in nodes of olfactory and high-order networks, and (v) modulation of regional cerebral glucose metabolism by the intensity and the duration of SARS-CoV-2-related loss of smell.

Included patients were mainly young adults (mean age: 42.6 years) with a very low frequency of comorbidities (e.g., hypertension, diabetes) usually associated with severe forms of COVID-19. Their main symptoms were altered sense of smell and taste; the latter being reported by all but one patient. Objective assessment of taste was not available. Considering the recognized ambiguity between taste and smell perception,[23,24] we prefered not relying on subjective assessment of taste and we concentrated the objective clinical assessment on the smell deficit. About 50-67% of the patients also had clinical signs of cold with cough, rhinorrhea or fever. Only one patient was hospitalized in a non-intensive COVID-19 ward for the management of his respiratory distress while all the other were outpatients. Overall, the clinical characteristics of our patients’ population are in line with those of SARS-CoV-2-positive patients with loss of smell and taste as predominant symptoms.[25–28] They contrast (in terms of age, comorbidities, clinical presentation) with those of patients with severe forms of COVID-19 who died in the same institution (i.e., CUB Hôpital Erasme) during the same period.[5]

The evolution of dysosmia was favourable in all patients but 58% of them still complained of incomplete recovery more than 15 weeks after the onset of symptoms. Long-lasting dysosmia in these patients could be the secondary consequence of the death of OE support and stem cells; the latter playing a crucial role in regenerating OE.[1]

### Structural MRI abnormalities

Bilateral (partial or complete) obliteration of the olfactory cleft was observed in about half of dysosmic patients. Other patients had either unilateral partial obliteration (three patients) or no MRI sign of olfactory cleft inflammation (four patients, one with signs of sinusitis). These findings are in line with imaging studies of the nasal cavity in COVID-19 patients with sudden loss of smell that either showed normal (see, e.g., [26,27,29]) or focal inflammation of the OE with obliteration of the olfactory cleft (see, e.g., [26,30]). This demonstrates that obliteration of the olfactory cleft due to focal OE inflammation is not the only possible pathophysiological mechanism underlying SARS-CoV-2-related dysosmia.[1]

Three dysosmic patients showed subtle OB asymmetry, with no OB MRI signal abnormality. One had complete and bilateral obliteration of the olfactory cleft, while the two others had either partial unilateral obliteration or no sign of OE inflammation at all. Previous MRI studies have demonstrated MRI changes in OB in patients with SARS-CoV-2-related anosmia[3,4,26] or in non-survivors of severe COVID-19,[5] suggesting some degree of central involvement at the origin of dysosmia in a subset of patients[1]. Results of the present study show that SARS-CoV-2-related OB involvement is not the main mechanism at the basis of acute loss of smell in COVID-19. Whether these OB abnormalities are associated with direct infection of OSNs or mitral cells remains doubtful considering the absence of ACE2 expression demonstrated in these cells.[1] Infection of vascular pericytes in the OB —in which there is evidence of ACE2 expression— might actually be responsible for OB MRI signal abnormalities found in some dysosmic patients. This hypothesis would be consistent with clinical evidence showing that SARS-CoV-2 predominantly influences the brain indirectly through effects on brain vasculature.[1]

No acute MRI signal abnormality was found downstream of the olfactory tract (or the rest of the brain), which is in line with most previous MRI studies performed in anosmic or severe forms of COVID-19 (see, e.g.,[3,5]). Still, rare cases of possible SARS-CoV-2-related limbic encephalitis have been reported in the literature (see, e.g., [31–33]), suggesting that SARS-CoV-2 might potentially reach the brain via, e.g., the olfactory nerves, like other neurotropic viruses such as herpes simplex virus.[34,35] Still, the question remains open since limbic encephalitis might be para-infectious (i.e., without local viral involvement) and the neuroinvasive capacity of SARS-CoV-2 to directly infect neurons remains, to the best of our knowledge, yet unproven.[1] MRI data from the present study do not support the involvement of central olfactory pathways in SARS-CoV-2-related sudden loss of smell.

Of note, a COVID-19 patient has been reported with FLAIR hyperintensities bilaterally in the OB and in the right gyrus rectus/orbitofrontal cortex four days after the onset of SARS-CoV-2-related anosmia.[3] In that case, FLAIR abnormalities resolved 28 days later on a subsequent MRI.[3] Based on this single case report, it might be speculated that OB and brain MRI abnormalities are only present at the very early phase of sudden loss of smell.[3] In this study, all PET-MR investigations were performed during the patients’ symptomatic phase, with 60% of them done >10 days after the onset of dysosmia. We cannot therefore exclude that some of our negative MRI findings are due to the rather late nasal and brain imaging. Further longitudinal studies are needed to better characterize the chronology of MRI abnormalities in this condition.

### Regional brain glucose metabolism abnormalities

At the individual level, heterogeneous (either decrease or increase) regional cerebral glucose metabolism abnormalities were found, in the core olfactory network and in other brain areas belonging to other sensory or high-level cognitive networks, as well as within the cerebellum. This finding highlights the absence of a clear and reproducible brain metabolic pattern across patients with SARS-CoV-2-related sudden loss of smell. These heterogeneous findings translate at the group-level into rather subtle (*p* < .001 uncorrected) regional decreases and increases in glucose consumption that, unexpectedly, did not mainly involve nodes of the core olfactory network *per se* (apart from the thalamus), but rather high-order neocortical areas. Although surprising at first glance, these individual- and group-level results are in line with (i) results of fMRI studies performed in healthy subjects showing that odor processing not only recruits the core olfactory network but also other brain structures such as high-order neocortical areas (for reviews, see, e.g., [16,36]), (ii) results of structural MRI investigations showing that dysosmic patients from various causes present changes in grey matter volume in numerous high-order neocortical areas similar to those observed in our population (see [37] for a review), and (iii) results of functional neuroimaging investigations done in dysosmic patients during olfactory stimulation, showing significant changes in the activity of similar high-order neocortical areas compared with healthy subjects.[20,37–39] Functional neuroimaging studies performed in humans have demonstrated that, while smelling single odors activates the core olfactory network, higher-level olfactory tasks (i.e., discrimination of odor quality, odor recognition memory or odor familiarity judgement) recruit brain areas outside the core olfactory network, including the cerebellum, cortices involved in other sensory modalities such as somatosensory or visual functions, and brain areas involved in high-order functions (for reviews, see [16,36,40]). The more complex the olfactory task is, the more remote these high-order areas are in terms of connexion with the core olfactory network.[16] These long-range connections explain part of the olfactory experience, i.e., odors can immediately elicit strong emotions, memories and mental images.[16] In light of this body of evidence, our results lead us to the hypothesis that SARS-CoV-2-related sudden loss of smell induces changes in glucose consumption in core olfactory and other brain areas due to alterations in both basic and higher olfactory functions.

Critically, correlations analyses between regional cerebral glucose metabolism and the severity/duration of SARS-CoV-2-related smell loss brought additional insights into the origin of the heterogeneity of the metabolic changes observed at the individual level. They indeed revealed a modulation of brain glucose consumption by the severity (negative correlation with the score on the “*identification test*”) and the duration (positive correlation) of dysosmia. Intriguingly, severe loss of smell (as documented by a low score on the “*identification test*”) was associated with a higher glucose metabolism in some olfactory core (e.g., orbitofrontal cortex) and high-order brain areas (FEF and PCC), while patients with more preserved olfactory function had lower glucose metabolism in these brain areas. Although these results appear rather counterintuitive, they are in line with those of a previous FDG-PET study, which also showed a negative correlation of regional glucose consumption with the level of loss of smell.[21] By contrast, longer duration of loss of smell was associated with a higher glucose metabolism in some core (e.g., orbitofrontal cortex) and high-order brain areas (DLPFC, MPFC, PCC). Importantly, no correlation was found between the intensity and the duration of smell loss. These correlation data shed light on the mixed, and somehow divergent, findings of individual- and group-level analyses on dysosmic patients. The regional decrease in metabolism, essentially observed in patients with moderate smell loss, is probably attributable to a deafferentation phenomenon as observed, e.g., in sudden visual loss due to non-infectious disorders.[41,42] By contrast, the increased or preserved metabolism, essentially associated with either severe or long-lasting loss of smell, could be related to functional reorganization/plasticity mechanisms. Such mechanisms involve, in particular, compensatory processes, unmasking of already existing but inactive neural connections, or the avoidance of maladaptive neural processes.[43] The hyperactivity of some brain areas may also relate to some forms of olfactory mental imaging associated with spontaneous repetitive odor search,[44] or to emotional/behavioral consequences of severe or prolonged anosmia.[45]

Considering that metabolic abnormalities were not associated with any MRI signal abnormality, they probably do not represent neuroimaging evidence supporting the neuroinvasive potential of the SARS-CoV-2 but rather functional cerebral markers of the olfactory deficit.

### Limitations of the study

PET-MR data were acquired in patients at different moments after the onset of dysosmia with no longitudinal neuroimaging follow-up. Although this approach proved its value in correlation analyses with regional cerebral metabolism, it did not allow proper characterization of the chronology of structural and metabolic abnormalities in COVID-19-related dysosmia.

This study did not include any objective evaluation of taste function as it was not available at our center at the start of the COVID-19 pandemic and was very difficult to obtain during this sanitary crisis. Considering that all but one patient complained about altered sense of taste, it would have been of great interest to investigate the brain metabolic changes specifically associated with dysgeusia. As discussed above, due to the recognized ambiguity between taste and smell perception,[23,24] we decided not to include any correlation analyses with the subjective evaluation of taste (i.e., visual analogue scale).

For voxel-based FDG-PET data analyses, we relied on a group of healthy subjects who had undergone PET-MR imaging in the context of an unrelated neuroimaging study. This was due to the difficulty to perform neuroimaging research investigations in healthy subjects in an academic hospital environment during the ongoing COVID-19 pandemics. For that reason, healthy subjects did not undergo any objective or subjective evaluation of smell and taste at the time of the PET-MR data acquisition. Also, the group of healthy subjects was not properly matched for age and sex with the included COVID-19 patients. Therefore, age was introduced as a covariate of non-interest in voxel-based FDG-PET data analyses, which limits the potential confound associated with this variable in our results. A similar approach could not be applied to accommodate the gender unbalance between patients and control groups, but the findings do not match reported gender differences in cerebral glucose metabolism [46–48], while being in line with results of previous FDG-PET studies on dysosmia.[9,20,21]

## Conclusions

This PET-MR study demonstrates that the main pathophysiological hypotheses (i.e., obliteration of the olfactory cleft, central involvement due to SARS-CoV-2 neuroinvasiveness) raised to explain sudden smell loss in COVID-19 do not explain dysosmia in all patients. It also shows that smell loss is associated with heterogeneous cerebral metabolic changes in core olfactory and other brain areas suggesting possible combined processes of deafferentation and active functional reorganization secondary to the lack of olfactive sensory stimulation. Based on those findings, SARS-CoV-2-infection limited to OE support and stem cells might be the predominant pathophysiological mechanisms involved in COVID-19 sudden loss of smell, with variable interindividual structural and functional consequences.

## Data Availability

De-identified brain MRI and FDG-PET data can be shared upon reasonable request for scientific purpose and after approval of the CUB Hopital Erasme Ethics Committee and authorities.

## Acknowledgments

The authors would like to express their most sincere gratitude to the PET-MR technologists who made this work possible, i.e., Maria Coelho De Vasconcelos, Ana Peralta Pereira, and Joao Marinho Ribeiro.

